# When Survival Improves But Quality of Life Does Not: A Model-Based Meta-Analysis of Immune Checkpoint Inhibitors

**DOI:** 10.64898/2026.03.04.26347610

**Authors:** Yimeng Sun, Shiwon Chang, Kayden Tang, Matthew R. LeBlanc, Adam C. Palmer, Malidi Ahamadi, Jiawei Zhou

## Abstract

**Background:** In immune checkpoint inhibitor (ICI) trials, overall survival (OS) benefits are well established, yet improvements in quality of life (QoL) are often inconsistent or absent in conventional analyses. This apparent discordance raises important questions: are QoL outcomes truly unrelated to survival, and how can QoL results be better utilized and interpreted?

**Methods:** A model-based meta-analysis (MBMA) of longitudinal EORTC QLQ-C30 global health status/quality of life data from randomized ICI trials was conducted. Longitudinal QoL trajectories were analyzed using a nonlinear mixed-effects model to estimate treatment-related toxicity and long-term QoL improvement. Associations between QoL trajectory parameters and OS were assessed using spearman rank correlation tests and Cox proportional hazards models.

**Results:** Twenty-seven studies (8,149 ICI and 5,593 control patients) contributed longitudinal QoL data, and 18 studies provided matched OS data. Raw QoL trajectories showed overlap between treatment arms, while OS consistently favored ICIs. MBMA revealed that ICIs had similar toxicity but significantly faster QoL improvement than control therapies (p < 0.0001). Baseline QoL, toxicity, and QoL improvement rate were all significantly associated with OS (p < 0.001). MBMA-based QoL comparisons were more sensitive in detecting associations with survival than raw QoL data, with the strongest association observed at Week 24 (R = -0.37, p = 0.067).

**Conclusions:** Conventional analyses comparing QoL at a single time point may obscure meaningful patient-reported benefits. By capturing longitudinal QoL trajectories across trials, MBMA reveals how patient experience evolves alongside survival outcomes and supports improved interpretation and utilization of QoL data in treatment evaluation.

## Introduction

Patient-reported outcomes (PROs), particularly health-related quality of life (QoL), play a critical role in ensuring that cancer treatments improve not only survival but also patients’ lived experience during and after therapy. [1, 2] Despite increasing inclusion of PROs in oncology trials, QoL endpoints are often underutilized in treatment evaluation and clinical decision-making. When QoL results appear inconsistent with overall survival (OS) findings, they are frequently viewed as “soft” evidence rather than central indicators of treatment benefit. [3, 4]

Immune checkpoint inhibitors (ICIs) exemplify this challenge. ICIs have revolutionized oncology by delivering durable overall survival (OS) benefits across a broad spectrum of solid tumors. However, across multiple randomized trials, including EORTC-18071, KEYNOTE-716, and IMpower150, ICIs significantly prolonged OS yet failed to demonstrate consistent or sustained improvements in QoL compared with control therapies. [5–7] This discrepancy raises an important question: are QoL outcomes truly unrelated to survival benefits in ICIs treatment?

As emphasized by the Setting International Standards in Analyzing Patient-Reported Outcomes and Quality of Life Endpoints Data (SISAQOL) initiative, inconsistent QoL and OS findings across trials are largely caused by the differences in how QoL endpoints are analyzed and interpreted. [8] Heterogeneity in trial design and statistical methodology, such as timing of assessments, handling of missing data, and selection of minimally clinically important difference thresholds, limit cross-trial comparability and reduce the clinical interpretability of QoL findings. [8–10] Traditional meta-analyses often compare the change from baseline in QoL between treatment arms at a single follow-up time point (e.g., the study endpoint) [11, 12], which can be limited by the heterogeneity in assessment schedules and follow-up durations across trials. Moreover, QoL changes over time in a dynamic and often nonlinear manner. Treatment efficacy and toxicity can have differential and time-dependent effects on OS and QoL (**Figure 1A**). As a result, clinically meaningful signals in QoL data may be obscured in the meta-analyses based on a single time point, limiting the ability of clinicians, regulators, and patients to fully understand the impact of treatment on daily life.

**Figure 1.**
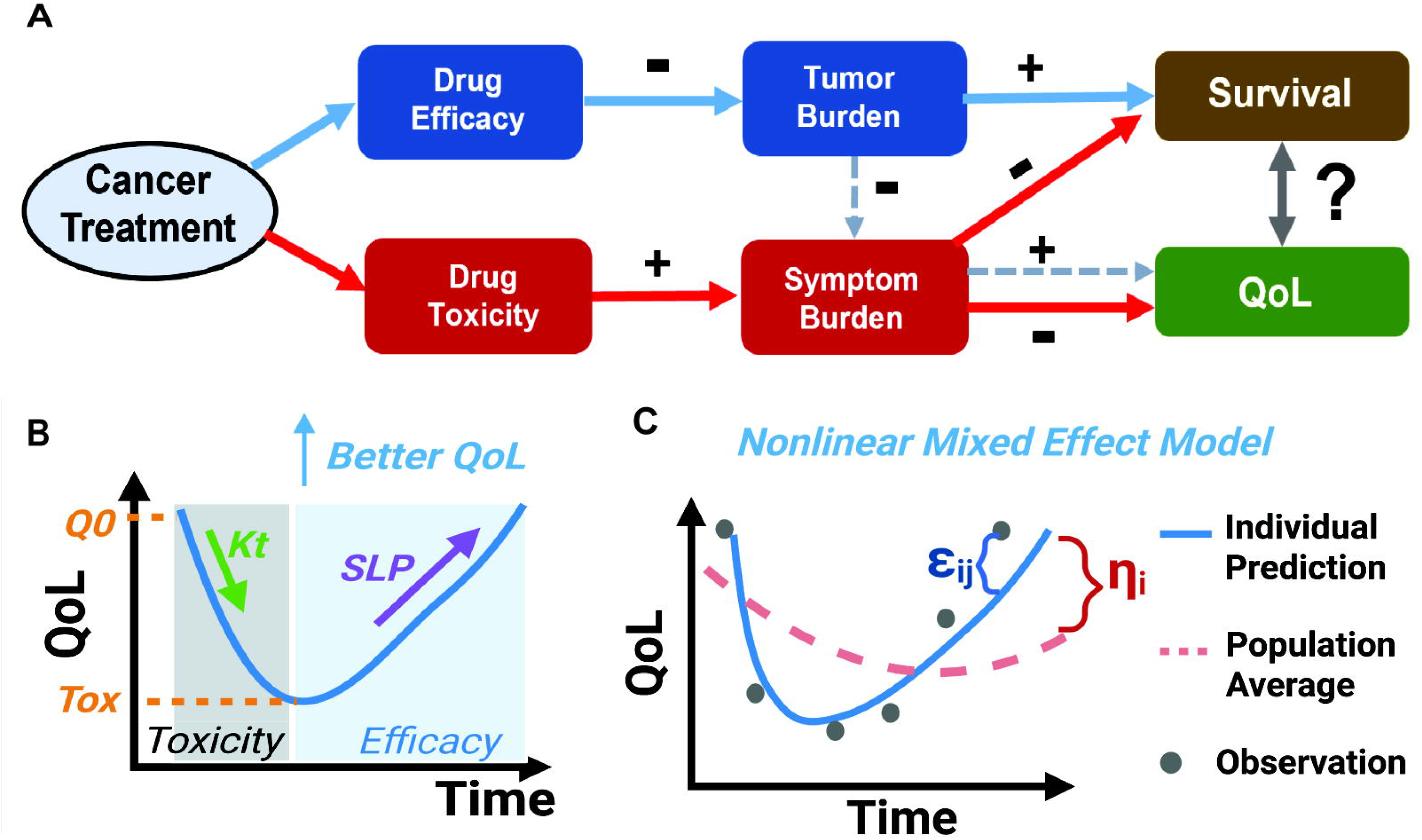
Study concept, model structure, and nonlinear mixed effect modeling approach used in the model-based meta-analysis. **(A)** Proposed relationships among treatment, efficacy and toxicity, and their associations with survival and quality of life (QoL). “+” represents an increase and “–” represents a decrease in the indicated outcome. **(B)** Semi-mechanistic model used to describe how QoL changes during treatment. The model reflects how drug efficacy and toxicity together influence patients’ QoL trajectories. **(C)** Diagram of nonlinear mixed effect modeling approach. All available longitudinal QoL data across studies are analyzed together in a single model. This approach estimates both the average trend in the population and the variability between studies or arms. The term *η_i_* represents how much an individual arm-level QoL trajectory differs from the population average; and ε_ij_ represents random variation with the same study (study ***i***) at measurement time point ***j***.

To better understand how patient experience evolves during cancer treatment, analytical approaches must capture the full trajectory of QoL over time. Model-based meta-analysis (MBMA) directly addresses this challenge. MBMA pools longitudinal QoL data across trials while accounting for differences in follow-up duration, assessment frequency, and sample size using nonlinear mixed effect (NLME) modeling approach. [13, 14] Importantly, this framework can separate short-term treatment-related toxicity from longer-term recovery or improvement in QoL, yielding clinically interpretable parameters that reflect distinct aspects of patient experience. [15] In our previous work, we successfully applied this approach to pembrolizumab trials and identified the QoL benefits that were obscured by noise and heterogeneity in traditional meta-analyses. [11, 15]

To examine the associations between survival and QoL, we conducted an MBMA of longitudinal EORTC QLQ-C30 global health status/quality of life data together with OS Kaplan–Meier data from randomized clinical trials of approved ICIs. By jointly modeling QoL trajectories across heterogeneous trial designs, we evaluated whether longitudinal QoL patterns are meaningfully associated with OS. We also examined whether our MBMA framework can overcome the analytical inconsistencies highlighted by the SISAQOL initiative. Through this approach, we aim to clarify how patient-reported QoL trajectories relate to survival outcomes and to provide a framework that better incorporates patient voice into the evaluation of cancer therapies.

## Methods

### Study design

This MBMA was prospectively registered in PROSPERO (CRD420251103645) and conducted in accordance with the Preferred Reporting Items for Systematic Reviews and Meta-Analyses (PRISMA) guidelines, as well as established guidance for MBMA. [16, 17]

### Search strategy and inclusion criteria

We conducted an electronic search of the PubMed database to identify clinical trials published up to June 12, 2025 that evaluated 11 approved ICIs (ipilimumab, pembrolizumab, nivolumab, atezolizumab, avelumab, durvalumab, cemiplimab, tremelimumab, retifanlimab, dostarlimab, and toripalimab). Studies were eligible for inclusion if they reported longitudinal European Organisation for Research and Treatment of Cancer Quality of Life Questionnaire–Core 30 (EORTC QLQ-C30) global health status/quality of life (GHS/QoL) scores (hereafter referred to as QoL scores) measured at three or more time points. QoL scores range from 0 to 100, with higher scores indicating better overall health status and QoL. [18] Studies that did not report longitudinal QoL data or reported QoL at fewer than three time points were excluded. For studies meeting the QoL inclusion criteria, OS Kaplan–Meier curves were additionally collected only when both ICI and control arm OS data were available. QoL and OS data were extracted from published figures using WebPlotDigitizer (https://automeris.io/) and digitizeit (www.digitizeit.xyz), respectively.

### Population QoL trajectory model

Longitudinal QoL trajectories were analyzed using MBMA approach, applying the same semi-mechanistic modeling framework as our previously published study. [15] QoL trajectory was described using a mechanistic nonlinear model that simultaneously captured two clinically distinct processes: an early, treatment-related decline reflecting toxicity, and a longer-term linear improvement reflecting recovery and overall well-being (**Figure 1B**, **Equation 1**). This mechanistic model allows separation of short-term toxicity from sustained QoL benefit, a key advantage over conventional time-point–based meta-analyses.

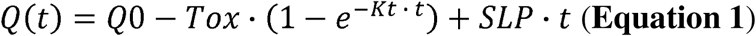

Where *Q(t)* is the QoL scores over time, *Q0* is the QoL score at baseline, *SLP* is the QoL improvement rate, *Tox* is the maximal toxicity, and *Kt* is the toxicity onsite rate, *t* is the time in week.

The key method in MBMA is to estimate the model parameters in **Equation 1** using an NLME modeling approach. NLME modeling is a statistical method that analyzes all available data from multiple individuals or studies at the same time within a single framework. [19] Instead of calculating results separately for each patient or each study and then averaging them, NLME fits one model to the entire dataset by estimating both the typical (average) pattern in the population and how much individuals differ from that average (**Figure 1C**). This approach is particularly suitable for clinical data with high variability, such as patient-reported QoL data. [19] In this study, it separates total variability into two components: between-study/arm variability, which reflects true differences among studies and treatment arms (**Equation 2**), and random variation, which reflects measurement error and within-study fluctuations over time (**Equation 3**). By explicitly modeling these sources of variability, the method prevents random noise from being misinterpreted as treatment effects.

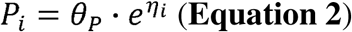

Where *P_i_* is the value for parameter *P* in study *i*, and *θ_P_* is the population average value for P, *η_i_* is the variability between study *i* and the population average (between-study/arm variability).

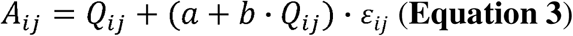

Where *A_ij_* is the observed QoL scores for study *i* at time *j*, Q_ij_ is the model predicted QoL. E_ij_ represents the random variation for study *i* at time *j* (within-study variability). The *a* and *b* are the parameters to be estimated in the model.

NLME model parameters were estimated using the stochastic approximation expectation–maximization algorithm. [20] Treatment effects (ICI vs control) were evaluated as covariates on key model parameters governing baseline QoL, treatment-related toxicity ***Tox***, and longitudinal QoL improvement rate ***SLP***. Additional baseline covariates (including age, sex, ECOG performance status, ICI type, and cancer type) were explored based on clinical relevance and data availability using a full model estimation approach. [21] Model adequacy was assessed using standard goodness-of-fit diagnostics, visual predictive checks, and evaluation of parameter precision. [22] Model validation was performed using nonparametric bootstrap resampling 500 times at the treatment-arm level to assess robustness and quantify parameter uncertainty.

### Statistical analysis for QoL and OS associations

Wilcoxon tests were applied to compare QoL trajectory parameters between ICIs and control arms. Associations between QoL trajectory parameters and OS outcomes were evaluated using Spearman rank correlation tests and Cox proportional hazards models. All statistical tests were two-sided. A p value <0.05 was considered statistically significant.

### Software

The population QoL trajectory model was developed in Monolix 2024R1 and the model simulations were performed using Simulx 2024R1 (https://lixoft.com/products/). Plots and statistical analyses were performed using R 4.4.1 and RStudio Version 2022.07.1+554 and compiled using Adobe Illustrator 2026.

## Results

Among 2,221 publications reporting QoL outcomes for 11 ICIs, 27 studies met the eligibility criteria by providing longitudinal EORTC QLQ-C30 QoL data after removal of duplicates (**Figure 2**). Of these, 18 studies also reported corresponding OS Kaplan–Meier curves that could be matched to the QoL data. In total, 8,149 patients in ICI arms with 316 longitudinal QoL assessments and 5,593 patients in control arms with 206 longitudinal QoL assessments were included in the QoL analyses, while 6,301 patients in ICI arms and 4,202 patients in control arms were included in the OS analyses. The final set of studies with both QoL and OS data comprised 25 between-arm comparisons, of which 20 reported evaluable results for both QoL and OS benefits. Study characteristics are summarized in **Table 1**.

**Figure 2.**
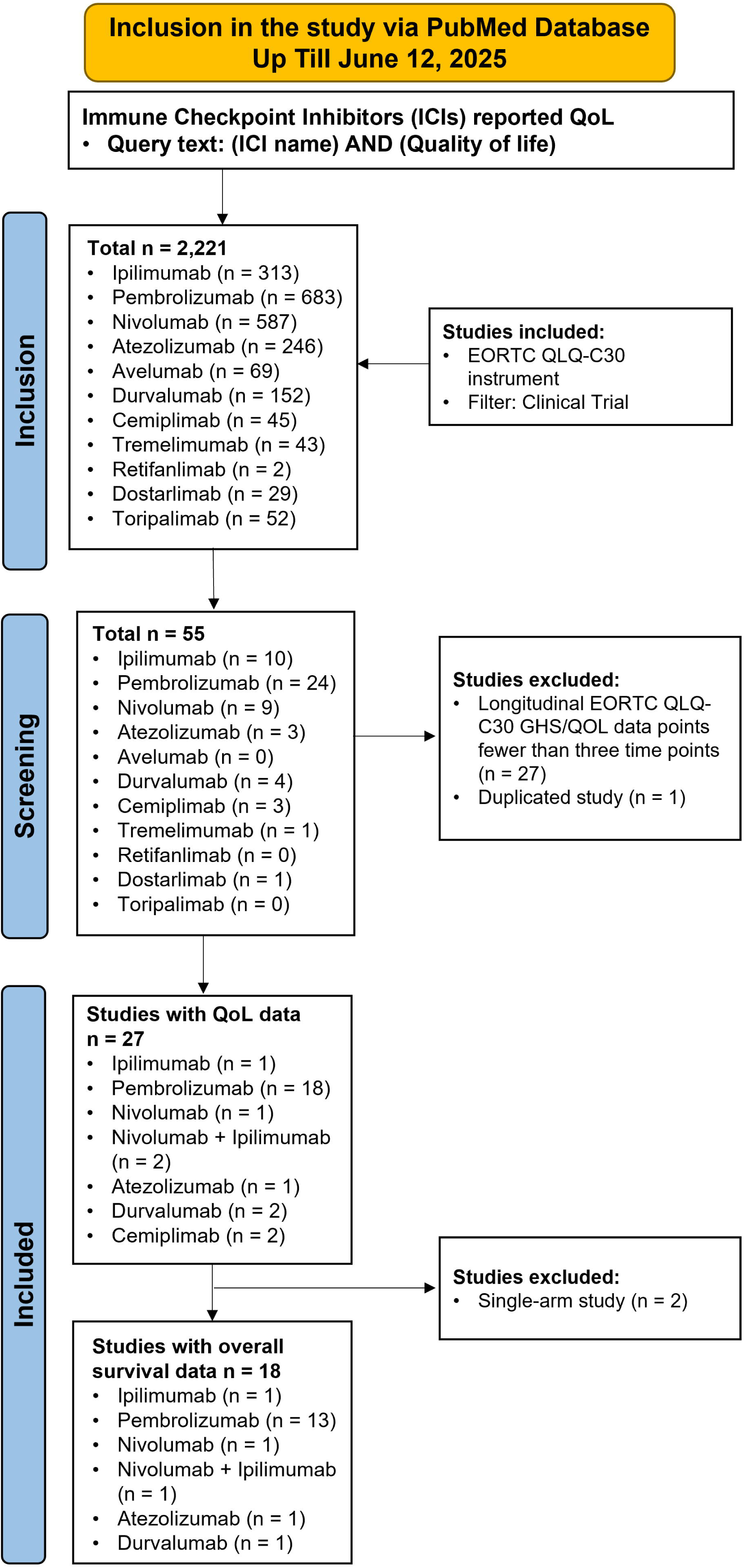
Study inclusion and exclusion criteria. A total of 27 studies reporting longitudinal EORTC QLQ-C30 GHS/QOL scores were included. Corresponding study-level overall survival Kaplan–Meier curve data were also collected when available.

**Table 1.**
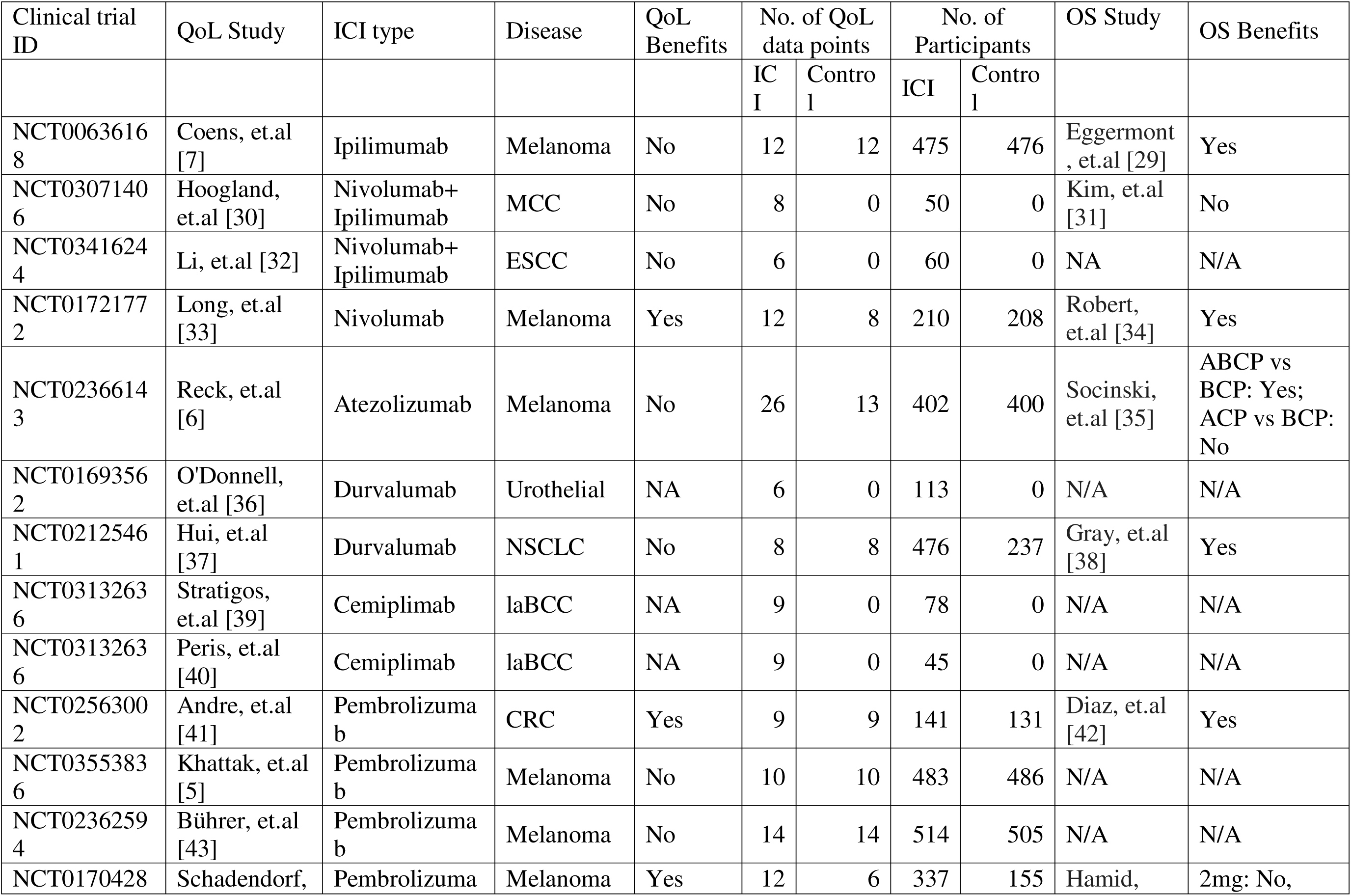
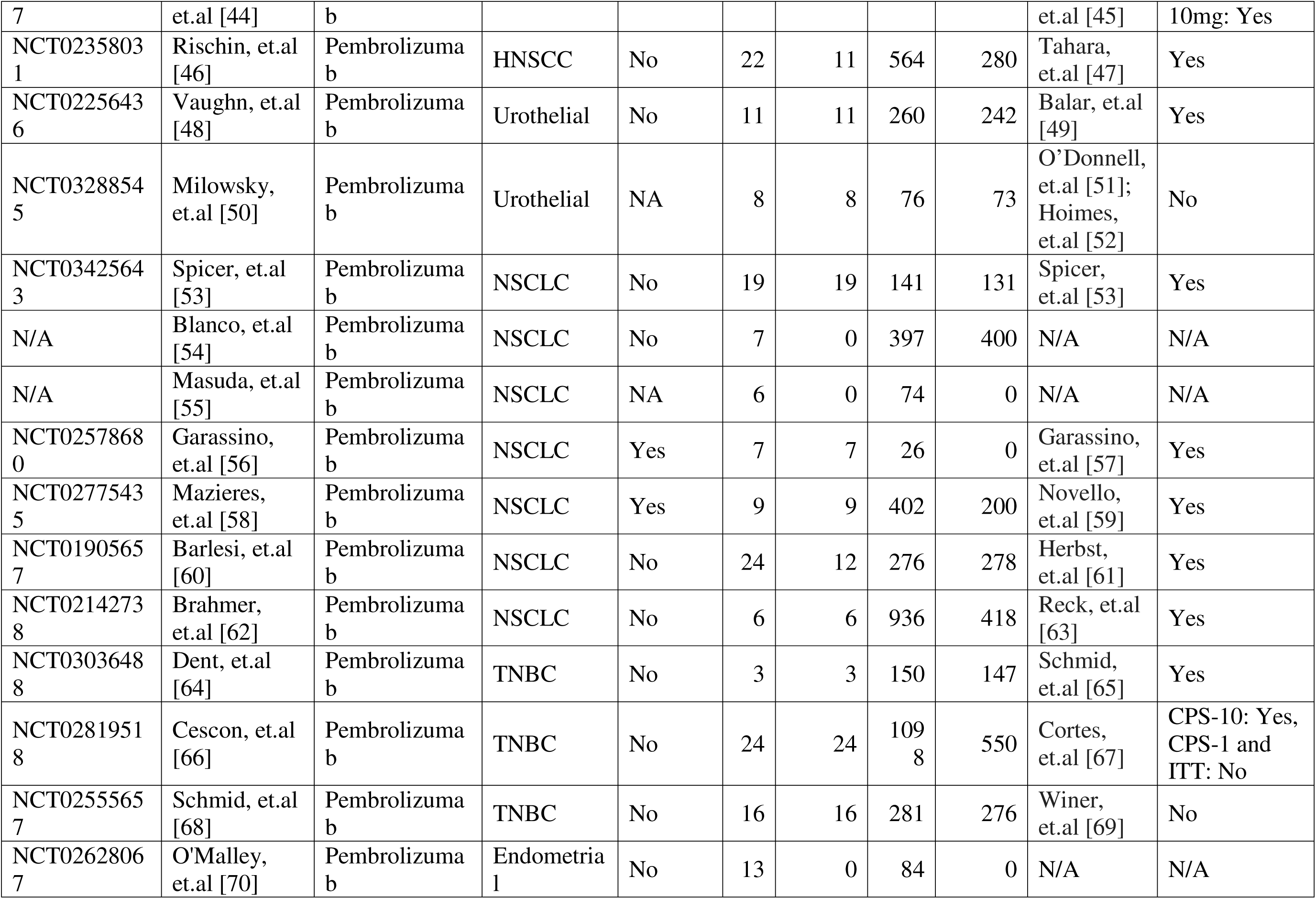
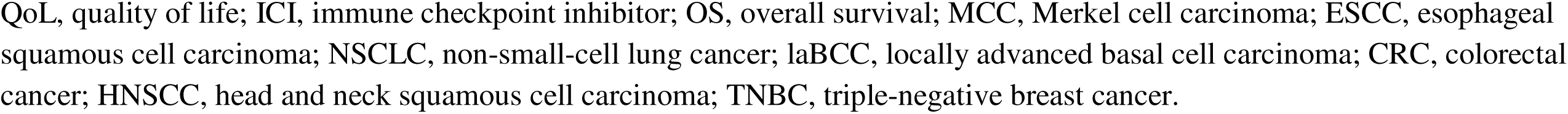
Summary of Study Information.

### Inconsistent observed QoL and OS benefits with ICIs

Longitudinal digitized QoL data showed substantial heterogeneity across trials, with largely overlapping trajectories between ICI and control arms and no clear evidence of QoL benefits favoring ICIs in raw data (**Figure 3A**). Assessment of publication bias using funnel plots showed no clear asymmetry; however, larger studies tended to demonstrate minimal differences in QoL between treatment groups, whereas smaller studies showed greater variability and more frequent apparent QoL benefit with ICIs (**Figure 3B**). In contrast, digitized Kaplan–Meier curves demonstrated consistent and durable OS benefits favoring ICIs across the majority of included trials (**Figure 3C**). Arm-level mapping indicated frequent discordance between QoL and OS outcomes, with statistically significant OS prolongation often observed in ICI arms despite no statistically or clinically meaningful improvement in QoL endpoints (**Figure 3D**).

**Figure 3.**
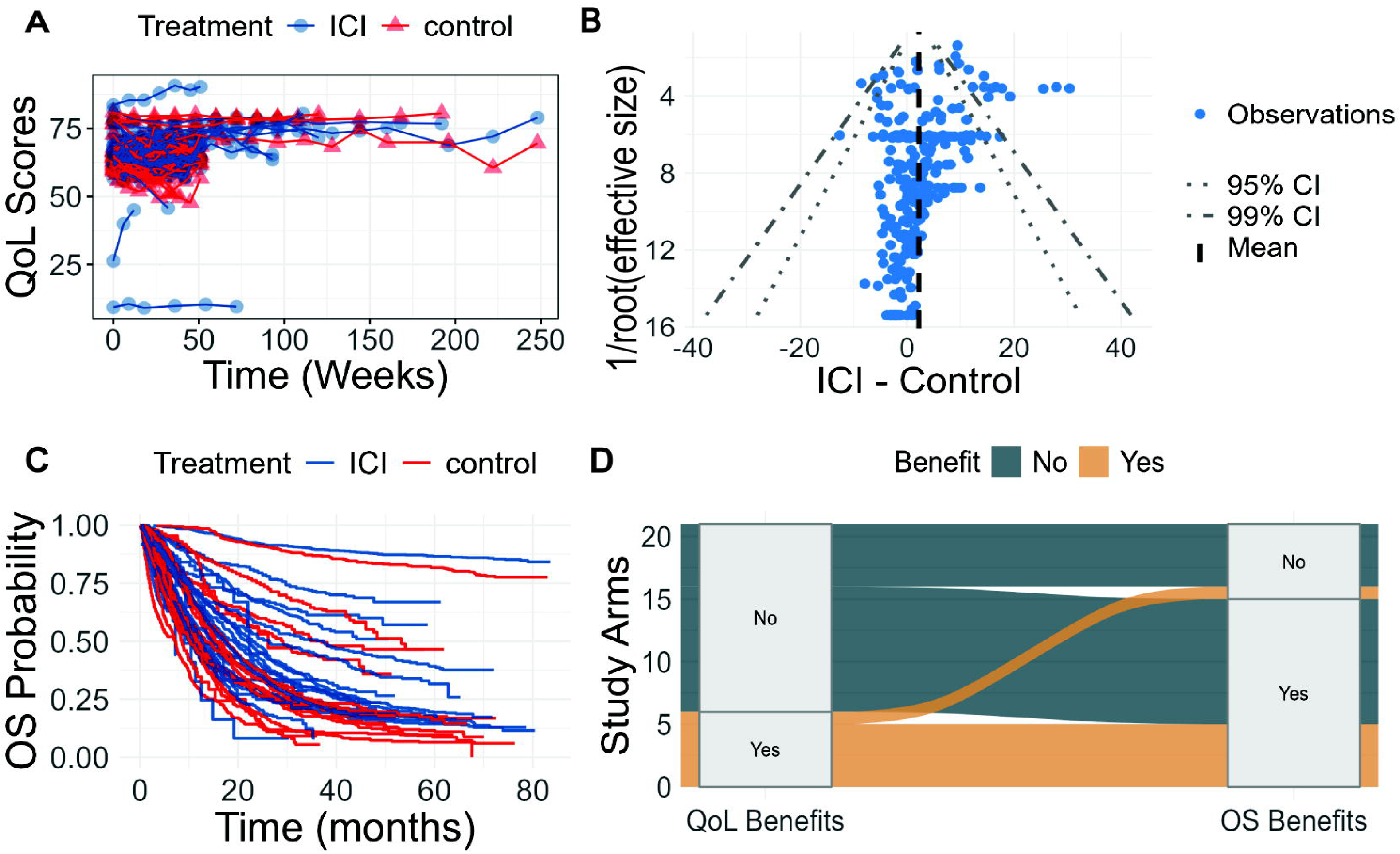
Patient-reported quality of life (QoL) and overall survival (OS) data collected in the study. **(A)** Digitized QoL scores change over time in immune checkpoint inhibitor (ICI) and control arms. Each circle represents an arm-level aggregated data point, and lines connect repeated measurements within the same arm across time. **(B)** Funnel plot showing the difference in QoL scores between ICI and control arms plotted against the inverse square root of the effective sample size (reverse of ICI arm sample size minus reverse of control arm sample size). The solid black line indicates the mean effect size, and the 95% and 99% confidence intervals are shown with different dashed line styles. **(C)** Digitized Kaplan–Meier curves for OS in ICI and control arms, obtained from the same clinical trials from which QoL data were collected. **(D)** Sankey plot of how clinical trials map between QoL benefit categories (statistically and/or clinically significant vs not significant) and OS benefit categories (statistically significant vs not), with flow widths proportional to the number of trials in each combination.

### MBMA identifies clinically interpretable QoL benefits and associations with OS

A NLME QoL trajectory model was developed using arm-level longitudinal QoL data, while handling between-study and between-arm variability. The model adequately captured longitudinal QoL data across studies, with good agreement between model predictions and observed QoL scores at both the population and individual levels. Diagnostic assessments indicated no major systematic bias, with well-behaved residuals over time and across predicted values. (**Figure 4A**) Visual predictive checks demonstrated that the model reproduced the central tendency and variability of QoL trajectories over follow-up (**Figure 4B**). Study-level goodness-of-fit analyses showed that predicted trajectories closely aligned with observed QoL patterns across individual trials, supporting the model’s ability to transform heterogeneous and noisy QoL data into clinically interpretable parameters. (**Figure 4C**) Model parameter estimates are summarized in **Table S1** and were estimated with good precision, with bootstrap validation confirming the robustness and stability of the model.

**Figure 4.**
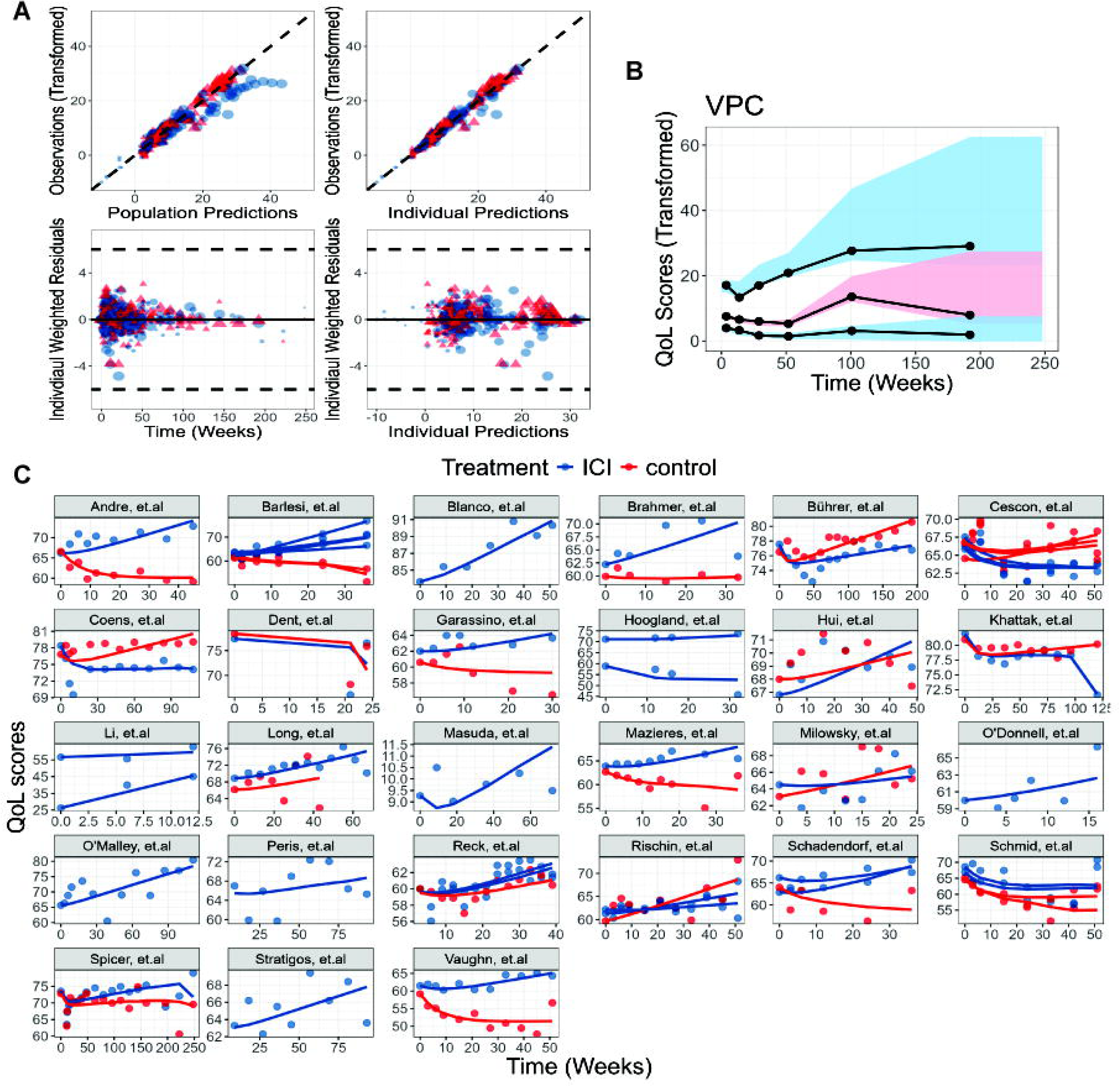
Population QoL trajectory model well captured the data. **(A)** Diagnostic plots showing population predictions versus transformed observations (top left) and individual predictions versus transformed observations (top right), with the identity line (y = x) shown as a black dashed line. Residual diagnostics include individual weighted residuals versus time (bottom left) and versus individual predictions (bottom right), with a solid black line at y = 0 and dashed lines at y = ±6. Red circles denote pembrolizumab arms and blue triangles denote control arms; symbol size reflects arm-level sample size. **(B)** Visual predictive check (VPC) of transformed quality of life (QoL) scores over time across all studies. Observed medians and 10th/90th percentiles are shown as black solid lines, while model predictions from 500 simulations are shown as shaded areas: red for the 90% prediction interval (PI) of the median and blue for the 90% PI of the 10th and 90th percentiles. **(C)** Study-level goodness-of-fit. Each panel corresponds to an individual study, with dots representing observed QoL scores and solid lines representing model-predicted individual QoL trajectories. ICI, immune checkpoint inhibitors.

Noisy patient-reported QoL data were transformed into clinically interpretable QoL trajectory parameters through population modeling, including baseline QoL, treatment-related toxicity (***Tox***), and QoL improvement rate (***SLP***). These parameters reflect different aspects of patient experience during treatment, including baseline well-being, treatment-related symptom burden, and the rate of recovery or improvement over time. ICIs showed similar baseline QoL and toxicity compared with control arms (p > 0.05; **Figure 5A-B**). However, ICIs had significantly faster QoL improve rate than control arms (p < 0.0001; **Figure 5C**), demonstrating a clear treatment benefit. These parameters were also associated with OS. Baseline QoL was significantly correlated with 12-month OS probability (correlation coefficient R = 0.45, p = 0.0015; **Figure 5D**). In contrast, ***Tox*** and ***SLP*** were not significantly correlated with 12-month OS probability in univariable correlation analyses (**Figure 5E–F**); however, both parameters showed significant associations with OS in the Cox proportional hazards model. Specifically, higher treatment-related toxicity was associated with worse OS (p < 0.001), whereas a higher QoL improvement rate was associated with better OS (p < 0.001; **Figure 5G**).

**Figure 5.**
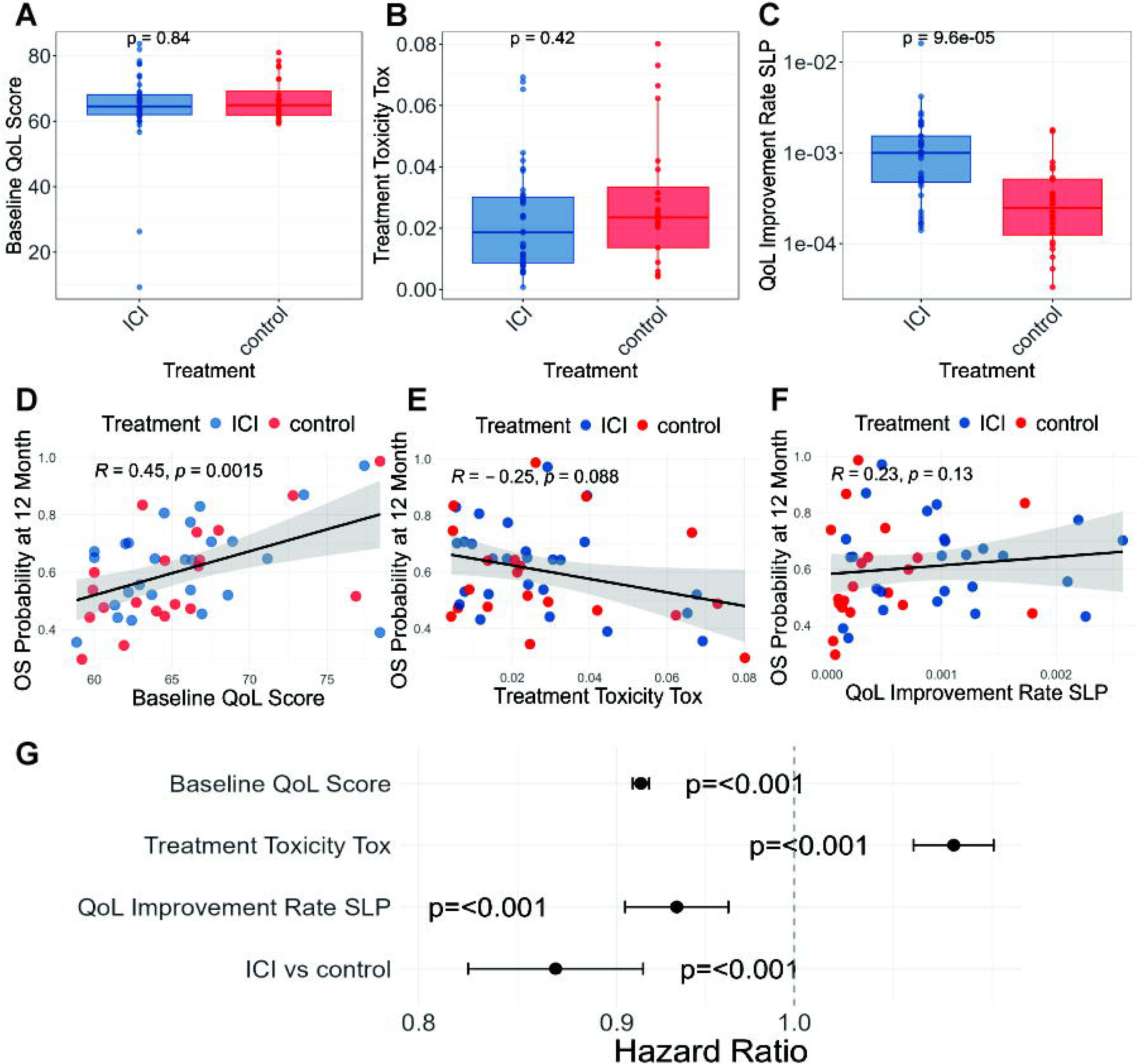
Overall survival (OS) is associated with quality of life (QoL) trajectory parameters. **(A-C)** Wilcoxon tests comparing baseline QoL **(A)**, treatment-related toxicity *Tox* **(B)**, and the QoL improvement rate *SLP* **(C)** between ICIs and control arms. **(D-F)** Linear regressions relating 12-month OS probability to baseline QoL **(D)**, treatment-related toxicity *Tox* **(E)**, and the QoL improvement rate *SLP* **(F)**. Each circle represents a study arm. **(G)** Forest plot of the cox proportional hazards model evaluating the association between QoL trajectory parameters and OS, adjusted for treatment type. ICI, immune checkpoint inhibitors.

### Association between ICI-Control Arm QoL differences and OS Hazard Ratio

Using observed between-treatment differences in QoL, correlations with OS hazard ratios were consistently negative but did not reach statistical significance at any evaluated time point (p>0.2), largely because QoL assessments were conducted at different time points across trials, limiting comparability (**Figure 6A**). In contrast, analyses based on model-predicted between-treatment QoL differences demonstrated stronger and more consistent associations with OS across time points, with the strongest association observed at Week 24 (R = -0.37, p = 0.067; **Figure 6B**). These findings suggest that MBMA incorporates studies with different follow-up durations into a unified analytical framework, enabling inclusion of a greater number of trials and supporting a more robust and comprehensive evaluation of treatment effects.

**Figure 6.**
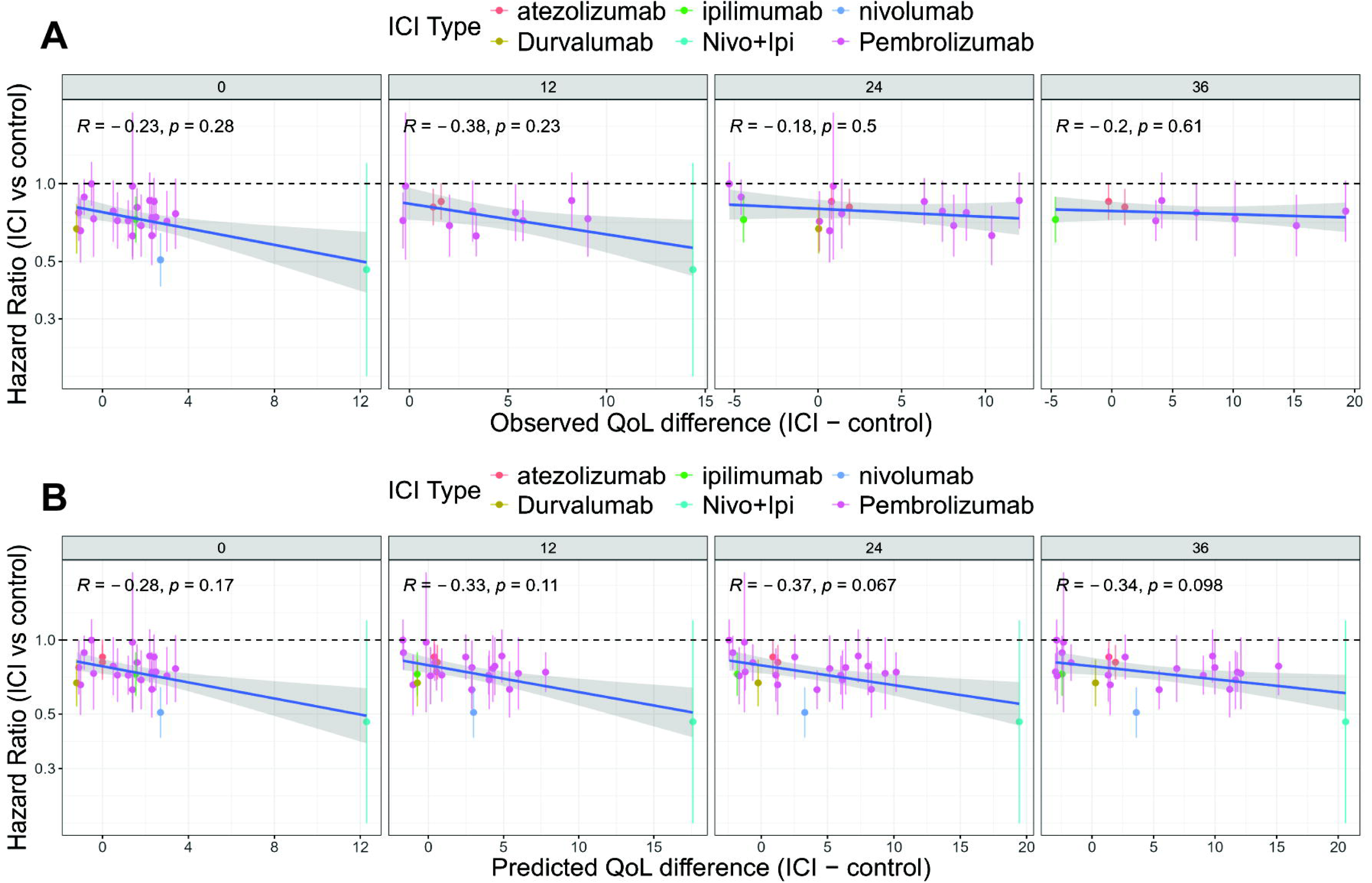
Association between treatment-related quality of life (QoL) differences and overall survival (OS). Observed between-treatment differences in QoL scores **(A)** or model-predicted between-treatment differences in QoL scores **(B)** at 0, 12, 24, and 36 weeks are plotted against the corresponding between-treatment OS hazard ratios.

## Discussion

In this study, we aim to better understand how patient-reported QoL change over time during ICI treatments. By analyzing longitudinal QoL data from multiple randomized studies, we found that although conventional single time-point analyses often show little difference in QoL between treatment arms, modeling longitudinal trajectories revealed clearer treatment-related patterns.

Specifically, ICIs demonstrated similar treatment-related QoL deterioration compared with control therapies but showed a significantly faster rate of QoL improvement over time. In addition, baseline QoL, treatment-related toxicity, and the rate of QoL improvement were all associated with overall survival.

At the study level, QoL data are largely overlapped between treatment arms but survival benefits with ICIs were more consistently observed. (**Figure 3A** and **Figure 3C**) This disconnect likely reflects limitations in how QoL data are analyzed rather than a true absence of patient benefit. [19] QoL measures are inherently variable, and missing data are common in oncology trials. Differences in assessment schedules, follow-up duration, and statistical methods further complicate QoL comparisons across studies. [8–10] As a result, clinically meaningful changes in patient-reported QoL may be difficult to detect using conventional analyses, which typically evaluate QoL by comparing the change from baseline between treatment arms at a single follow-up time point.

Our findings directly address these challenges. By pooling longitudinal QoL trajectories and accounting for differences in follow-up, assessment frequency, and sample size across trials, MBMA provides a unified framework for synthesizing QoL data. This approach preserves more information from longitudinal patient-reported outcomes and enables more consistent interpretation of QoL results across studies.

Importantly, the absence of a clear QoL improvement does not necessarily mean a lack of benefit. [23, 24] When ICIs prolong OS, maintaining stable QoL itself represents a meaningful clinical success. Improved survival without QoL deterioration suggests that patients live longer without compromising their well-being. [23, 24] However, this pattern may be difficult to detect using raw QoL scores with cross-trial comparisons, which can obscure clinically relevant longitudinal QoL effects. [15] From the patient perspective, this raises concern that important aspects of treatment experience may be overlooked when QoL data are analyzed by single time point. In this study, we used an MBMA approach to better uncover these relationships. By separating variability in QoL data into between-arm differences and within-study random variation, we were able to clarify the treatment effects, showing comparable toxicity profiles but a greater rate of QoL improvement with ICIs.

MBMA is a quantitative method within the broader framework of model-informed drug development (MIDD), which uses modeling and simulation to integrate diverse data and inform regulatory decision-making. [25] MIDD has been shown to improve drug development efficiency; for example, a Pfizer report estimated that MIDD saved approximately 5 months in development time and $0.5 million cost per drug on average. [26] As a MIDD tool, MBMA enables structured cross-trial synthesis of heterogeneous data and translates variable patient-reported QoL data into clinically interpretable QoL trajectory parameters linked to survival. This approach provides a more integrated evaluation of patient-centered efficacy and toxicity, supports quantitative benefit–risk assessment, and may enhance the regulatory value of QoL endpoints in oncology.

Several limitations should be acknowledged. This analysis relied on arm-level summary data rather than individual patient-level data, which limits assessment of within-patient variability. QoL and OS data were incompletely reported across trials, reducing the number of studies available for joint analysis. Moreover, the observed associations between QoL trajectories and survival are exploratory and should not be interpreted as causal. In addition, both OS and QoL outcomes may vary by tumor type and line of therapy, which were not fully accounted for in this pooled analysis and could influence the observed results.

Another potential source of error was the low image quality of published figures, where rasterization and overlapping features may have slightly affected data extraction. [27, 28] However, digitized traces reproduced nearly all segments of the published curves with high fidelity. Any minor discrepancies fall within the intrinsic confidence intervals of the Kaplan–Meier estimator and are unlikely to meaningfully affect the results.

Taken together, these findings support a more integrated role for patient-reported QoL in evaluating the benefits of ICIs and potentially other cancer therapies. By examining longitudinal PROs across trials, this framework helps reveal meaningful patterns in patient experience that may be overlooked in conventional analyses. Improving how QoL data analyzed and interpreted will improve the utilization of PROs in oncology trials and help ensure that treatment evaluation more fully reflects the perspectives and experiences of patients.

## Acknowledgements

The authors thank the patients and their families and caregivers and investigators for participating in the studies used in this analysis. We acknowledged the use of AI tools (OpenAI, ChatGPT) to improve the language in the manuscript writing.

## Data availability statement

Source data and codes are available at https://github.com/zhoujw14/MBMA_ICI.git.

