## Supplementary Materials for "When Survival Improves But Quality of Life Does Not: A Model-Based Meta-Analysis of Immune Checkpoint Inhibitors"

**Table S1. Final QoL trajectory model parameter estimates.**

| **Parameter** | **Value** | **RSE (%)** | **Bootstrap median** | **Bootstrap 10^th^/90^th^ Percentile** | **SHR (%)** |
| --- | --- | --- | --- | --- | --- |
| **Population parameters** | | | | | |
| Maximal toxicity reducing QoL (*Tox*) | 0.0146 | 26.8 | 0.0154 | 0.008 – 0.024 |  |
| Toxicity onset rate (*Kt*, 1/week) | 0.134 | 22.9 | 0.143 | 0.071 – 0.27 |  |
| QoL improvement rate (*SLP*, 1/week) | 6.09×10^-4^ | 26.6 | 6.86×10^-4^ | 4.67×10^-4^ – 1.05×10^-3^ |  |
| Effect of control arm on *SLP,* ICI as reference ($\theta_{SLP\_PBO})$ | -1.1 | 44.8 | -1.2 | -1.98 – -0.47 |  |
| Between-study variability on *Tox* (*BSVTox*) | 0 Fixed | | | | |
| Between-arm variability on *Tox* (*BTATox*) | 0 Fixed | | | | |
| **Inter-individual variability (standard deviation)** | | | | | |
| Standard deviation of between-study variability on *Tox* ($\Omega_{BSVTox}$) | 0.87 | 23 | 0.91 | 0.49 – 1.43 | 23.5 |
| Standard deviation of between-arm variability on *Tox* ($\Omega_{BTATox}$) | 5.49 | 22.4 | 5.96 | 2.19 – 10.9 | 14.5 |
| Standard deviation of between-study variability on *SLP* ($\Omega_{SLP}$) | 1.21 | 14.1 | 1.06 | 0.77 – 1.36 | 13.1 |
| **Correlations** | | | | | |
| Correlation between *Tox* and *SLP* | -0.853 | 38.4 | -0.82 | -0.88 – -0.54 |  |
| **Residual unexplained variability** | | | | | |
| Additive residual error (***a***) | 0.557 | 9.51 | 9.52 | 0.47 – 0.73 |  |
| Proportional error (***b***) | 0.0627 | 11.5 | 0.058 | 0.025 – 0.083 |  |

RSE, relative standard error; CI, confidence interval; SHR, shrinkage.

**Source Model Codes.**

A. **Monolix codes.**

```
 1. DESCRIPTION:
 2. MBMA for QOL.
 3.
 4. [LONGITUDINAL]
 5. input = {EmaxFE, Kp, SLP, BSL, NOC, etaBSVEmax, etaBTAVEmax }
 6. NOC    = {use=regressor}
 7. BSL    = {use=regressor}
 8.
 9.
10. EQUATION:
11.
12.
13. ; Maximal QoL (improved)
14. ; transform the Emax fixed effect (EmaxFE) to tEmax (normally distribute)
15. tEmax = logit(EmaxFE)
16.
17. ; adding the random effects (RE) due to between-study variability and
18. tEmaxRE = tEmax + etaBSVEmax +  etaBTAVEmax/sqrt(NOC)
19.
20. ; transforming back to have EmaxRE with logit distribution (values between 0 and 1)
21. Emax = exp(tEmaxRE)/(1+exp(tEmaxRE))
22.
23.
24. ; Define QoL trajectory
25. SLP2 = SLP*0.001
26. Q = BSL*0.01 - Emax * (1 - exp(-Kp * t)) + SLP2*t
27. ; adding a saturation to avoid taking logit(0) (undefined) when t=0
28. Qsat = min(max(Q,0.01),0.99)
29. Qscore = Qsat*100
30.
31. ; transforming the effect Q in the same way as the data
32. pred = logit(Q)*sqrt(NOC)
33.
34. OUTPUT:
35. output = {pred}
36. table ={Qscore, Emax, SLP2}
37.
```

B. **Monolix settings.**

```
 1. <DATAFILE>
 2.
 3. [FILEINFO]
 4. file={path='../Data_MBMA_Monolix_Clean_10.24.2025.csv'}
 5. delimiter = comma
 6. header={ID, STID, OID, PUBMEDID, NCT, DISEASE, DRUG, DRUG2, TIMEWK, TransDV, RESPONSEFINAL, NTOTAL, NOC, BSL, AGE, Male, ECOG0, ECOG1, DSI, DSII, DSIII, DSIV}
 7.
 8. [CONTENT]
 9. STID = {use=identifier}
10. OID = {use=occasion}
11. PUBMEDID = {use=covariate, type=categorical}
12. DISEASE = {use=covariate, type=categorical}
13. DRUG = {use=covariate, type=categorical}
14. DRUG2 = {use=covariate, type=categorical}
15. TIMEWK = {use=time}
16. TransDV = {use=observation, type=continuous}
17. NOC = {use=regressor}
18. BSL = {use=regressor}
19.
20. <MODEL>
21.
22. [COVARIATE]
23. input = {DISEASE, DRUG, DRUG2, PUBMEDID}
24.
25. DISEASE = {type=categorical, categories={'CRC', 'ESCC', 'Endometrial', 'HNSCC', 'MCC', 'Melanoma', 'NSCLC', 'TNBC', 'Urothelial', 'laBCC'}}
26. DRUG = {type=categorical, categories={'Cemiplimab', 'Durvalumab', 'Nivo+Ipi', 'Pembrolizumab', 'atezolizumab', 'control', 'ipilimumab', 'nivolumab'}}
27. DRUG2 = {type=categorical, categories={'ICI', 'control'}}
28. PUBMEDID = {type=categorical, categories={'27405322', '27596353', '28162999', '29129441', '29590008', '30711649', '31581306', '31601496', '31751163', '32035514', '32459597', '33812497', '35381576', '35488720', '35835611', '36202690', '37557022', '37976633', '38070159', '38215355', '38913881', '39021272', '39031963', '39073799', '39097500', '39146951', '39288781'}}
29.
30. DEFINITION:
31. DISEASE2 =
32. {
33.   transform = DISEASE,
34.   categories = {
35.   'G_Melanoma' = {'Melanoma'},
36.   'G_NSCLC' = {'NSCLC'},
37.   'G_TNBC' = {'TNBC'},
38.   'G_CRC_ESCC_Endometrial_HNSCC_MCC_Urothelial_laBCC' = {'CRC', 'ESCC', 'Endometrial', 'HNSCC', 'MCC', 'Urothelial', 'laBCC'}  },
39.   reference = 'G_CRC_ESCC_Endometrial_HNSCC_MCC_Urothelial_laBCC'
40. }
41.
42. [INDIVIDUAL]
43. input = {EmaxFE_pop, Kp_pop, SLP_pop, etaBSVEmax_pop, omega_etaBSVEmax, gamma_SLP, etaBTAVEmax_pop, gamma_etaBTAVEmax, DRUG2, beta_SLP_DRUG2_control, corr2_etaBTAVEmax_SLP}
44.
45. DRUG2 = {type=categorical, categories={'ICI', 'control'}}
46.
47. DEFINITION:
48. EmaxFE = {distribution=logitNormal, typical=EmaxFE_pop, no-variability}
49. Kp = {distribution=logNormal, typical=Kp_pop, no-variability}
50. SLP = {distribution=logNormal, typical=SLP_pop, covariate=DRUG2, coefficient={0, beta_SLP_DRUG2_control}, varlevel=id*occ, sd=gamma_SLP}
51. etaBSVEmax = {distribution=normal, typical=etaBSVEmax_pop, sd=omega_etaBSVEmax}
52. etaBTAVEmax = {distribution=normal, typical=etaBTAVEmax_pop, varlevel=id*occ, sd=gamma_etaBTAVEmax}
53. correlation = {level=id*occ, r(etaBTAVEmax, SLP)=corr2_etaBTAVEmax_SLP}
54.
55. [LONGITUDINAL]
56. input = {a, b}
57.
58. file = 'test8.txt'
59.
60. DEFINITION:
61. TransDV = {distribution=normal, prediction=pred, errorModel=combined1(a, b)}
62.
63. <FIT>
64. data = 'TransDV'
65. model = TransDV
66.
67. <PARAMETER>
68. EmaxFE_pop = {value=0.01, method=MLE}
69. Kp_pop = {value=0.2, method=MLE}
70. SLP_pop = {value=0.7, method=MLE}
71. a = {value=1, method=MLE}
72. b = {value=0.3, method=MLE}
73. beta_SLP_DRUG2_control = {value=0, method=MLE}
74. c = {value=1, method=FIXED}
75. corr2_etaBTAVEmax_SLP = {value=0, method=MLE}
76. etaBSVEmax_pop = {value=0, method=FIXED}
77. etaBTAVEmax_pop = {value=0, method=FIXED}
78. gamma_SLP = {value=1, method=MLE}
79. gamma_etaBTAVEmax = {value=1, method=MLE}
80. omega_etaBSVEmax = {value=1, method=MLE}
81.
82. <MONOLIX>
83.
84. [TASKS]
85. populationParameters()
86. individualParameters(method = {conditionalMean, conditionalMode })
87. fim(method = Linearization)
88. logLikelihood(method = Linearization)
89.
90. [PLOTS]
91. run = true
92. plots = {indfits = {selected = true}, parameterdistribution = {selected = true}, obspred = {selected = true}, covariancemodeldiagnosis = {selected = true}, covariatemodeldiagnosis = {selected = true}, vpc = {selected = true}, residualsscatter = {selected = true}, residualsdistribution = {selected = true}, randomeffects = {selected = true}, saemresults = {selected = true}}
93.
94. [SETTINGS]
95. GLOBAL:
96. exportpath = 'test8'
97.
```
